# The Development and Evaluation of AI-based Tuberculosis Screening with a Digital Stethoscope used to Capture Lung Sounds. A Case-Control Study

**DOI:** 10.1101/2025.07.31.25332442

**Authors:** Max Rath, Johan Coetzee, Mark van Breda, Braden van Breda

## Abstract

**Background:** Tuberculosis (TB) remains a leading global cause of preventable death, with 10.8 million cases and 1.3 million deaths reported in 2023. Current methods for TB screening include symptom-based screening, and chest X-ray (CXR) with computer-aided detection (CAD-CXR). Each method has limitations related to cost, accessibility, and screening efficacy. As a result, an estimated 2.6 million TB cases were missed in 2023. AI-based TB screening using lung sounds captured by a digital stethoscope offers a potential solution to these challenges, enhancing access, efficacy, and cost-efficiency.

**Methods:** A dataset comprising 49,770 anonymized chest auscultation recordings from 1,659 participants (cases and controls) were collected by trained nurses in South Africa’s Western Cape province from June 2021 to November 2022 using AI Diagnostics’ prototype digital stethoscope. Consenting participants suspected to have TB that reported a recent sputum TB Xpert Ultra test were recruited from 34 primary care clinics. After stratification and data preparation, a final dataset of 1,169 participants was partitioned into an 80% training and 20% hold-out test set. A pre-trained transformer-based architecture was fine-tuned using K-fold cross-validation. The ensemble model’s ability to predict pulmonary TB was evaluated on the hold-out test set, with sputum Xpert Ultra as the reference standard.

**Results:** The AI model achieved a mean Area under the Receiver Operating Curve (AUC-ROC) of 0.79 (95% CI: 0.73-0.85). At a sensitivity of 89.9% (95% CI: 82.4%-94.4%), the ensemble model has a specificity of 50.4% (95% CI: 42.0%–58.7%) for predicting pulmonary TB using lung sounds.

**Conclusion:** AI-based digital chest auscultation for TB, with a sensitivity of 89.9% and specificity of 50.4% in this study, shows early promise as an alternative or adjunct to current TB screening methods. In addition, the method’s portability and low cost have the potential to significantly improve TB screening access. Future independent studies in diverse, unselected populations with high TB prevalence are required to validate model generalizability.

**Key messages:** 

**What is already known on this topic:** Tuberculosis (TB) is a major global health issue, especially in low-resource areas. Existing screening methods like symptom checks, chest X-rays, and CAD tools are often costly, hard to access, or not sensitive enough. AI has shown promise in detecting other lung conditions using sound, but its use for TB screening has not been well studied.

**What this study adds:** This is the first large study showing early promise that AI can detect TB from lung sounds using a digital stethoscope. This technology could be further developed as a low-cost and portable screening tool which aligns well with the World Health Organization’s End TB Strategy.

**How this study might affect research, practice, or policy:** This study would encourage further research into AI-based auscultation in different populations and settings, helping build more the evidence base supporting the use of AI in disease screening. Further research would also support the development of more accurate and generalizable models. In clinical practice AI-based digital stethoscopes could be used for early TB screening, allowing faster diagnosis and treatment. This would be especially important in asymptomatic TB cases where symptom-based screening would miss all cases. From a policy perspective, the results of this study would support further research which may support the inclusion of this technology in national and global TB screening guidelines and WHO endorsement.

This study was commercially funded by the technology provider, AI Diagnostics Pty (Ltd).

## Introduction

### Background

Tuberculosis (TB) is a leading cause of death globally, despite being preventable and curable. In 2023, 10.8 million people developed TB, and 1.3 million people died from TB. The WHO’s End TB Strategy places screening and early diagnosis as a top priority to end the global TB epidemic by 2035.^1^

The current WHO TB screening guideline includes symptom-based screening, chest X-ray (CXR) interpretation by a radiologist, and computer-aided detection software for CXR (CAD- CXR).^2^ According to the 2024 WHO target product profiles (TPPs) for TB diagnostics, a triage or screening test should achieve a minimum sensitivity of 90%, to minimize false negatives, and a minimum specificity of 70%.^3^

Current TB screening methods have numerous limitations: symptom-based screening has low sensitivity (71%) and specificity (64%), chest X-ray (CXR) interpretation is often limited by clinician availability and experience, and CAD-CXR remains costly and inaccessible in many low-resource settings.^2^ In addition, there is growing awareness of the need to conduct active case finding, since passive case finding often misses asymptomatic TB.^4^ As a result of the current issues with TB screening, an estimated 2.6 million cases were missed in 2023.^5^

Medical practitioners use chest auscultation to examine for abnormal lung sounds that aid in the diagnosis of pulmonary TB. The screening and diagnostic utility of normal and abnormal lung sounds depend on the expertise of the medical practitioner, which is developed over years of training and clinical experience. Access to formally qualified medical practitioners is low globally. This is more pronounced in low- and middle-income nations, where TB prevalence is often higher.^6^

Over the last decade, numerous studies have shown the potential of various machine learning (ML) approaches for lung sound classification. The classification performance of these models has been tested for diseases such as chronic obstructive pulmonary disease (COPD), asthma, and pneumonia.^7^ In a 2023 review of 62 studies on ML for abnormal lung sound detection (ASD) and respiratory disease recognition (RDR), the accuracy of more advanced ML models for RDR was over 90%.^8^ This illustrates the significant potential of ML for the classification of various lung diseases using digital lung auscultation. There is currently little evidence on the performance of ML models for TB detection using lung sounds.

This study was conducted in the Western Cape, South Africa, in a population disproportionately affected by TB due to low socioeconomic status and limited access to healthcare.^9^ This TB burden is compounded by high HIV prevalence in the region which exacerbates health inequalities.^10^ Access to low cost, accurate TB screening methods in this and other high-burden populations could help reduce health disparities, improve early detection, reduce missed cases, and significantly reduce TB mortality.

### Objectives

This study aims to evaluate the performance of an artificial intelligence (AI) model trained on lung sound recordings captured via digital chest auscultation for the detection of pulmonary TB.

## Methods

### Study design

This case-control study was conducted to investigate the performance of an AI model for TB screening using lung sounds collected using digital chest auscultation. Only participants from primary care clinics with suspected TB were recruited to reflect a real-world screening setting. Cases included participants with recently microbiologically confirmed TB via sputum Xpert Ultra, while controls recruited from the same clinics included individuals suspected to have TB but with a recent negative Xpert Ultra result.

This case-control study defined sample size requirements based on statistical power calculations with a 1:1 case-control ratio and 5% type I error rate. For a hypothesized 90% sensitivity and 70% specificity, the calculated sample sizes needed for varying precision levels were: ±2.5% error margin (1,106 participants for sensitivity and 2,581 for specificity), ±5% error margin (277 participants for sensitivity and 645 for specificity), and ±10% error margin (69 participants for sensitivity and 161 participants for specificity). The study initially targeted 3,200 participants (1,218 cases, 1,982 controls) for 95% confidence intervals (CI) with a ±2.5% width. The final dataset included 1,169 participants (493 TB-positive and 676 TB-negative), falling within the required sample size target range for evaluating an early-phase AI-based screening approach. ^11^

Figure 1 shows the enrolment process, criteria, and numbers of participants and lung sounds collected. Informed consent was obtained from all participants. For participants under the age of 18, consent was obtained from a parent or legal guardian, and assent was obtained from the participant where appropriate.

**Figure 1:**
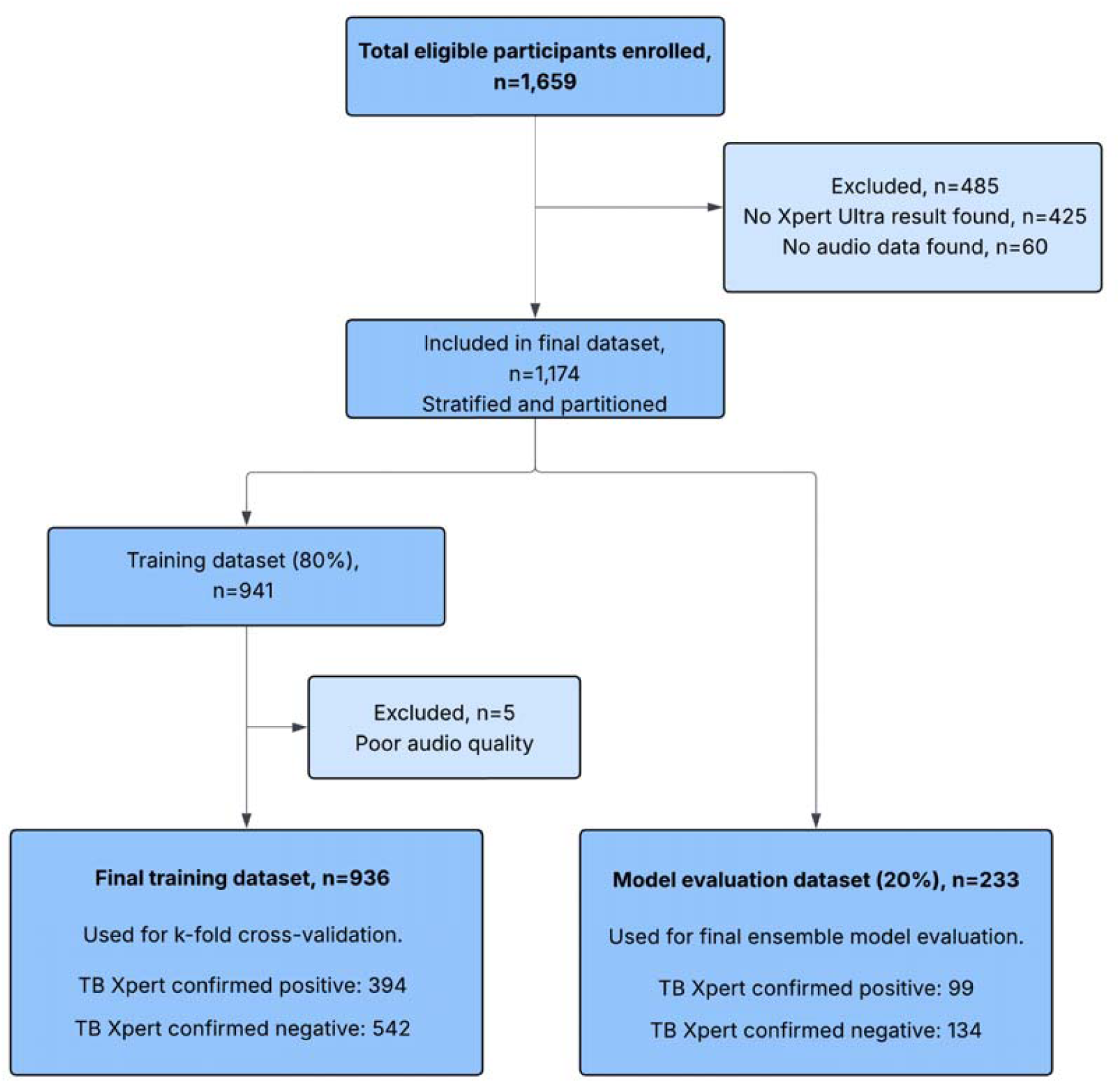
Outline of the participant inclusion and exclusion process used for model training and evaluation.

Patients and the public were not involved in the design, data collection, or analysis in this study.

### Data collection

In 2020 AI Diagnostics, a South African MedTech start-up, designed and built a simple prototype digital stethoscope for the recording of lung sounds. Using this prototype digital stethoscope, trained primary care nurses in the Western Cape, South Africa, captured an anonymized dataset of 49,770 chest auscultation recordings from consenting 1,659 participants (30 recordings per participants), male and female, aged 14 or above, between June 2021 and November 2022.

Consenting participants suspected to have TB that reported a recent sputum TB Xpert Ultra test were recruited from 34 primary care clinics. Participants were booked for chest auscultation data collection using the digital stethoscope. Nurses received training on use of the digital stethoscope prior to use. Nurses recorded at six auscultation positions (Figure 2) with participants’ chests exposed, with five breathing cycles (deep breaths, 3 seconds inspiration, 3 seconds expiration) per position. The six positions chosen minimized screening time while ensuring most of the chest surface area was covered. The entire data collection process took approximately five minutes per participant, was non-invasive, caused no discomfort and no harm to the participant or data collector.

**Figure 2:**
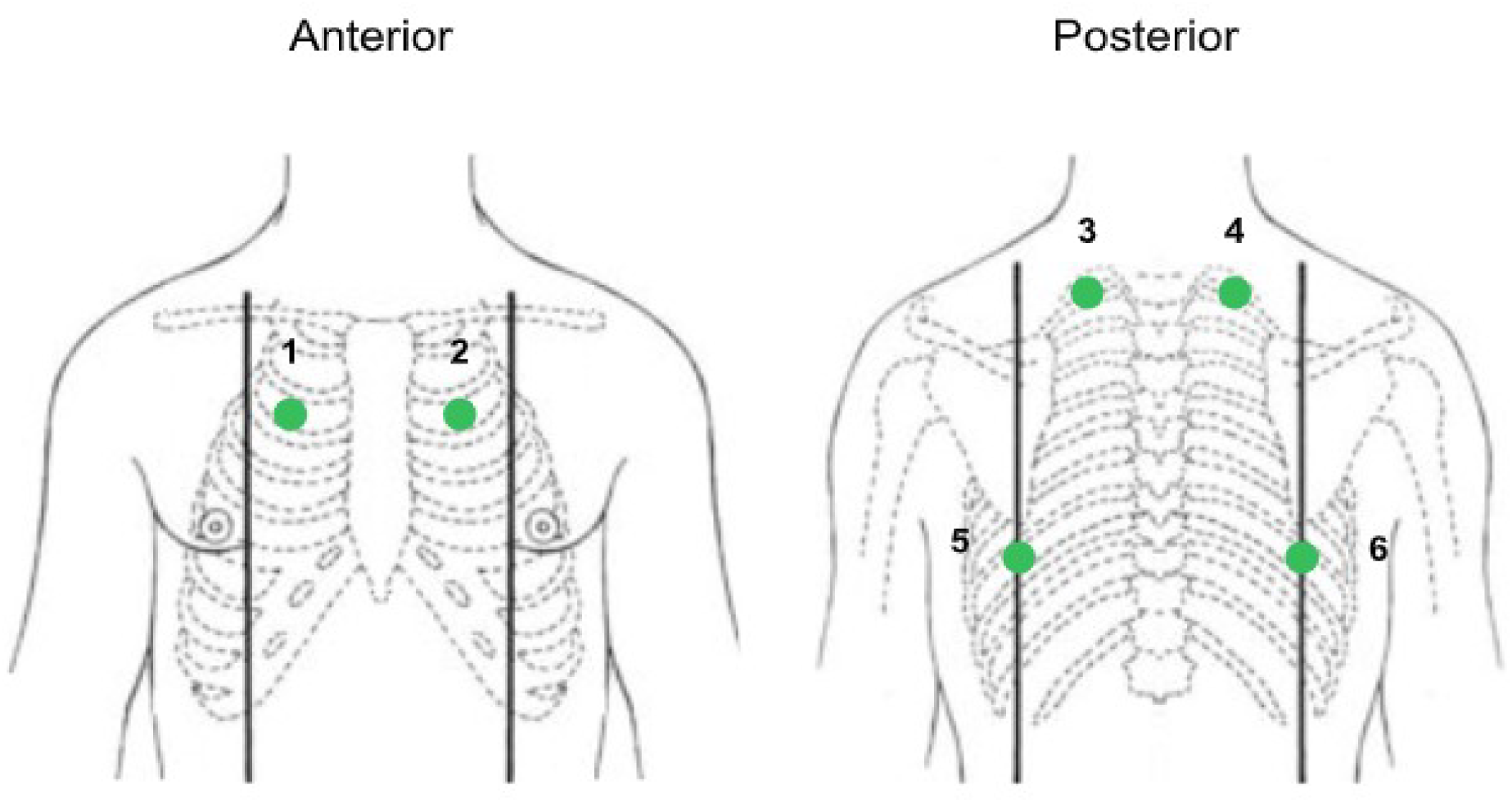
Six chest auscultation positions

Xpert Ultra results were checked for all participants included in the study. Demographic and clinical information was collected, as reported by each participant, and captured electronically prior to digital chest auscultation. Self-reported HIV status was not checked against any lab record of HIV status for each participant.

Participants were included in the training dataset only if a positive Xpert Ultra result was available within 14 days of data collection with the digital stethoscope, or if a negative Xpert Ultra result was available within 4 days of data collection. Trace positive Xpert Ultra results were also included in the dataset.

Of the 1,659 participants screened, 1,234 had an Xpert Ultra result within the designated timeframe. Only patients with a sputum Ultra result were included in the model evaluation to ensure a consistent and reliable reference standard to decide on presence or absence of TB (the ground truth). Xpert Ultra is a WHO-endorsed rapid molecular test with both high sensitivity (90.9%, 95% CI: 86.2%–94.7%) and high specificity (95.6%, 95% CI: 93.0–97.4%) for pulmonary TB, based on a Cochrane systematic review.^12^

### Data preparation

Python v3.11.9 was used for all data preparation and analysis. Graphing was performed using Matplotlib, Seaborn, and Graphs packages. Measures of statistical uncertainty were calculated using the confidence interval library. Data preparation was performed using scikit-learn. Deep learning models were created using PyTorch and trained with PyTorch. In preparation for model development and analysis, the study data set was stratified by the number of confirmed tuberculosis cases, HIV status, and symptom status. Unique participant identifiers were used for stratification at a participant level to ensure participant data fell into either the training set or test set and not split across both sets.

There were 60 participants removed from the dataset because the corresponding audio file was not found due to technical failure. The dataset was randomly partitioned in two: an 80% training set (n=941) and a hold-out 20% test set (n=233) to be used for external validation. The initial split reserves a portion of the data for final model evaluation that has not been seen during any part of the training process. Audio recordings with very poor audio quality (<10% usable audio, n=5) were removed from only the training dataset. The final dataset used for model training therefore included 936 participants.

No significant class imbalance was observed between TB positive and negative participants. Class sizes were monitored within each stratum to ensure interpretability of model performance. No balancing or reweighting was performed.

In the hold-out test set (n=233), there were 99 TB positive participants and 134 TB negative participants, 123 females and 110 males, 72 HIV positive participants and 161 HIV negative participants, 194 participants with at least one TB symptom and 39 asymptomatic participants.

### Model training

Raw audio data captured by the digital stethoscope was first transformed into a Mel spectrogram. This time-frequency representation of the audio data emphasizes the pitch range most relevant to human hearing and is frequently used in audio-based AI model development and evaluation.^7^ The model used Mel spectrograms as input into a sophisticated transformer-based deep learning architecture to generate an embedding of the audio. This embedding was then reduced to a binary output using linear neural network layers. The model was pre-trained and then fine-tuned on existing proprietary data. No recalibration was performed.

K-fold cross-validation was used in model training on the 80% split of the dataset. The training dataset was separated into 5 equal size folds (k=5). For each of the 5 iterations in the cross-validation, 4 folds are used for training a model, and the remaining fold is used for testing, such that each fold is used for validation. This process results in 5 slightly different models - AID.TB (1), (2), (3), (4) and (5). Each model was trained using the same method and hyperparameters.

The 5 models trained during the k-fold cross-validation were then combined to form a final ensemble model, AID.TB.

### Final ensemble model evaluation

The performance of this ensemble model was then evaluated using the initial 20% hold-out test set. The mean probability from the 5 models (AID.TB 1-5) was used assess the probability of pulmonary TB in each participant based on lung sounds and HIV status. This provided a final assessment of the model’s generalizability. An ROC curve was generated representing the model performance on the hold-out test set, accompanied by the corresponding AUC and 95% CI using the nonparametric percentage bootstrapping. The corresponding specificity at the WHO-targeted 90% sensitivity was included with a 95% CI using the Wilson exact score.

The threshold was selected on the validation set within each fold such that the sensitivity approximates the current WHO target of 90%. The corresponding specificity at that threshold was then derived. This method indirectly supports fairness, by reducing variability in false-negative rates, ensuring consistent assessment of model performance across strata. The same threshold was then used to calculate the sensitivity and specificity of the ensemble model on the hold-out test set at that threshold.

We did not account for heterogeneity in model parameters or performance between clinic sites. Standardized training and data collection protocols were used, and data collection performed by the same nursing team regardless of site. These measures minimized inter-site variability.

### Model Subgroup Evaluation

The model’s performance was further assessed on the hold-out test set in different key subgroups, namely HIV positive, HIV Negative, asymptomatic participants, and symptomatic participants. A symptomatic participant was defined using the South African national symptom-based screening guideline for TB, namely a participant with any one of: cough for more than 2 weeks, night sweats, weight-loss, and fever.^13^ Asymptomatic participants had none of these symptoms.

The AUC performance of the AID.TB ensemble model for each subgroup was determined using the 20% hold-out test set. The threshold was determined within each subgroup such that the sensitivity approximates the current WHO target of 90%. The specificity of AID.TB for each subgroup was then determined at that threshold. The Wilson exact score was again used for 95% CI.

## Results

### Study population characteristics

The baseline characteristics of the 1,169 participants in the dataset are detailed in **Table 1** below. There were 585 (51.3%) males and 569 (48.7%) females in the study population. The mean age was 37.1 (±13.3). Of the 1,169 participants in the study, almost a third, 363 (31.1%) were HIV positive, 519 (44.4%) were smokers, 65 (5.6%) had diabetes mellitus (type 1 or 2), and 55 (4.7%) had a history of asthma. There were 493 (42.2%) sputum TB Xpert Ultra positive participants in the study, and 676 (57.8%) sputum negative for TB. The high TB prevalence observed in the study was due to the case-control design and recruitment strategy employed.

**Table 1:**
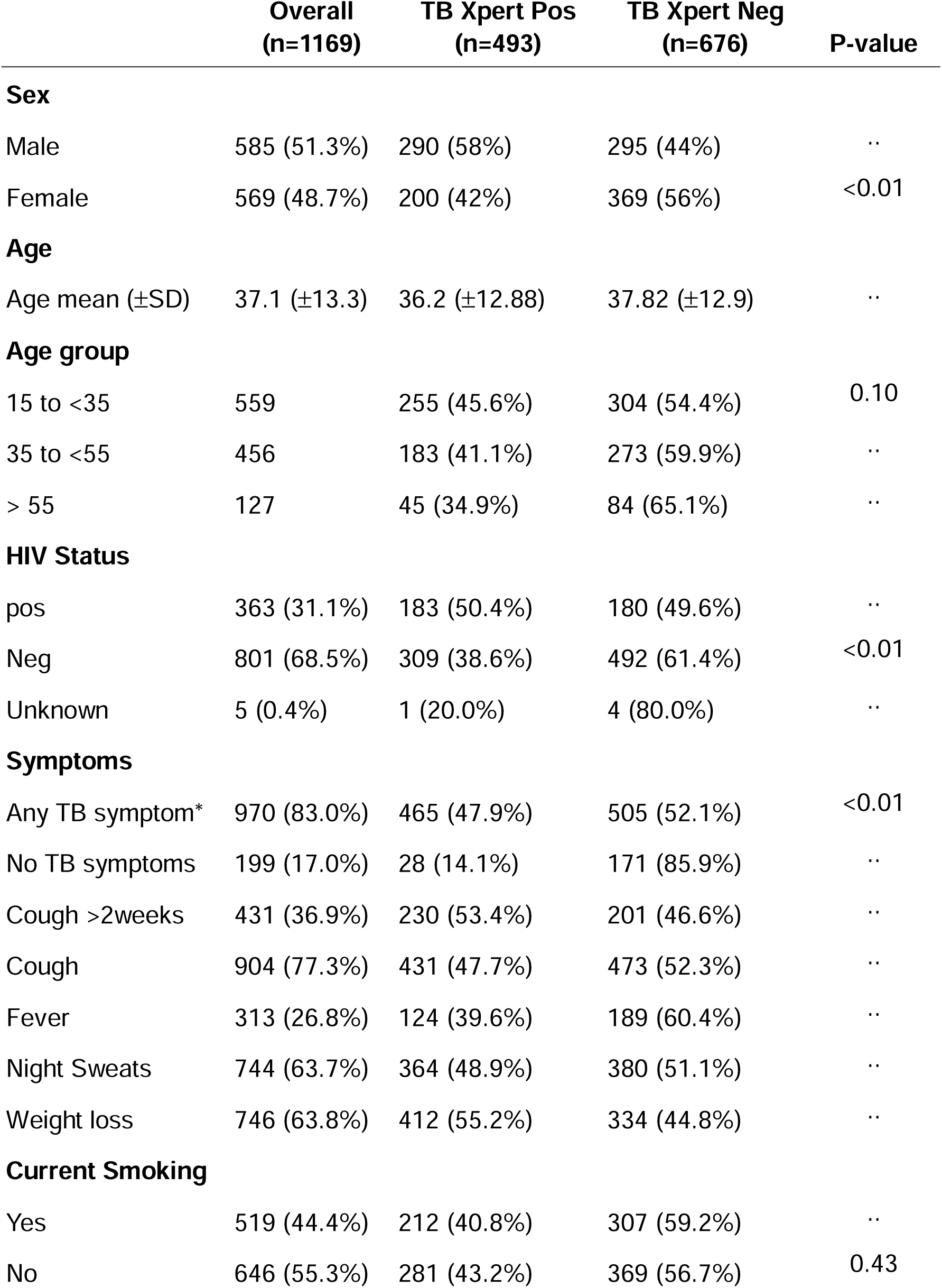
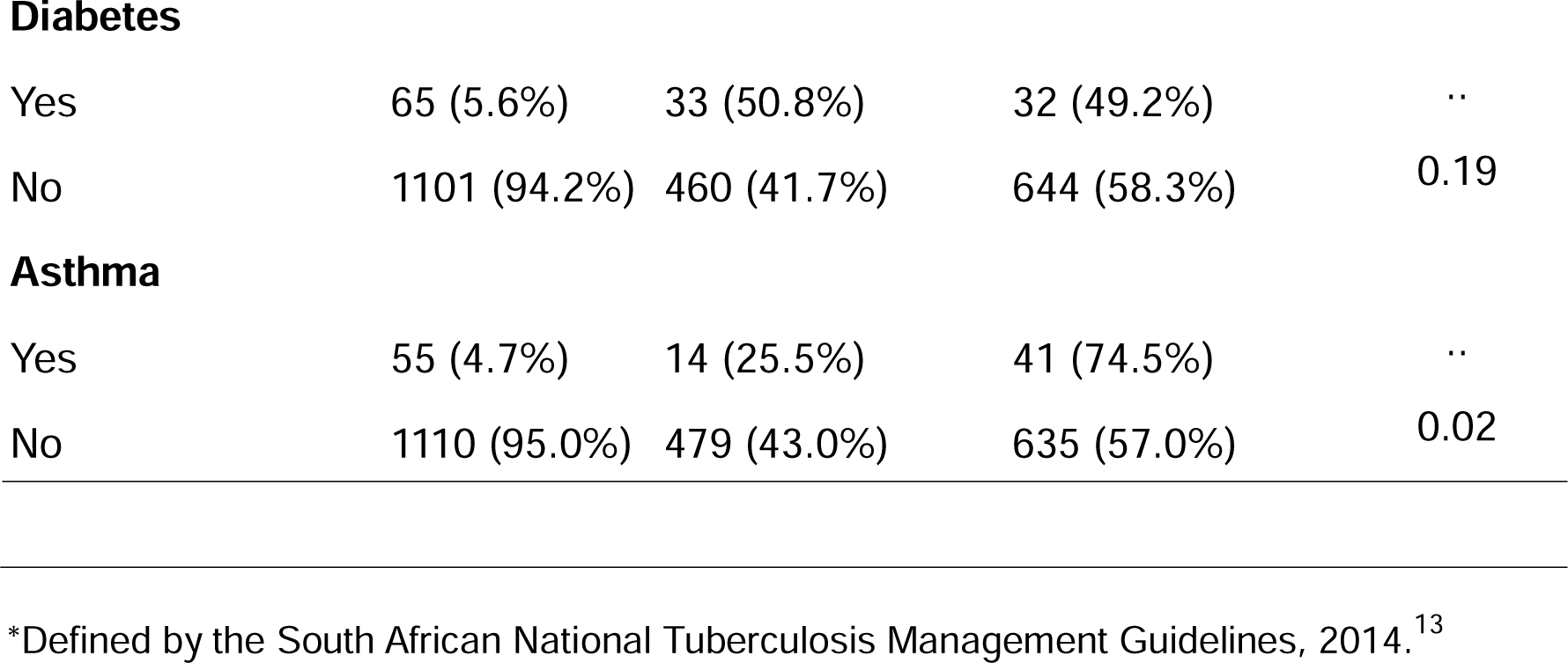
Baseline characteristics of the study population.

|  | **Overall**<br>**(n=1169)** | **TB Xpert Pos**<br>**(n=493)** | **TB Xpert Neg**<br>**(n=676)** | **P-value** |
| --- | --- | --- | --- | --- |
| **Sex** |  |  |  |  |
| Male | 585 (51.3%) | 290 (58%) | 295 (44%) | .. |
| Female | 569 (48.7%) | 200 (42%) | 369 (56%) | <0.01 |
| **Age** |  |  |  |  |
| Age mean (±SD) | 37.1 (±13.3) | 36.2 (±12.88) | 37.82 (±12.9) | .. |
| **Age group** |  |  |  |  |
| 15 to <35 | 559 | 255 (45.6%) | 304 (54.4%) | 0.10 |
| 35 to <55 | 456 | 183 (41.1%) | 273 (59.9%) | .. |
| > 55 | 127 | 45 (34.9%) | 84 (65.1%) | .. |
| **HIV Status** |  |  |  |  |
| pos | 363 (31.1%) | 183 (50.4%) | 180 (49.6%) | .. |
| Neg | 801 (68.5%) | 309 (38.6%) | 492 (61.4%) | <0.01 |
| Unknown | 5 (0.4%) | 1 (20.0%) | 4 (80.0%) | .. |
| **Symptoms** |  |  |  |  |
| Any TB symptom* | 970 (83.0%) | 465 (47.9%) | 505 (52.1%) | <0.01 |
| No TB symptoms | 199 (17.0%) | 28 (14.1%) | 171 (85.9%) | .. |
| Cough >2weeks | 431 (36.9%) | 230 (53.4%) | 201 (46.6%) | .. |
| Cough | 904 (77.3%) | 431 (47.7%) | 473 (52.3%) | .. |
| Fever | 313 (26.8%) | 124 (39.6%) | 189 (60.4%) | .. |
| Night Sweats | 744 (63.7%) | 364 (48.9%) | 380 (51.1%) | .. |
| Weight loss | 746 (63.8%) | 412 (55.2%) | 334 (44.8%) | .. |
| **Current Smoking** |  |  |  |  |
| Yes | 519 (44.4%) | 212 (40.8%) | 307 (59.2%) | .. |
| No | 646 (55.3%) | 281 (43.2%) | 369 (56.7%) | 0.43 |
| **Diabetes** |  |  |  |  |
| Yes | 65 (5.6%) | 33 (50.8%) | 32 (49.2%) | .. |
| No | 1101 (94.2%) | 460 (41.7%) | 644 (58.3%) | 0.19 |
| **Asthma** |  |  |  |  |
| Yes | 55 (4.7%) | 14 (25.5%) | 41 (74.5%) | .. |
| No | 1110 (95.0%) | 479 (43.0%) | 635 (57.0%) | 0.02 |

\*Defined by the South African National Tuberculosis Management Guidelines, 2014.<sup>13</sup>

Of the 199 participants in the study that had no TB symptoms on symptom-based screening, 28 (14.1%) were TB sputum positive. The median days between sputum collection and digital auscultation screening was 6 days for TB Xpert Ultra positive participants and 2 days for TB Xpert Ultra negative participants.

### Performance of AID.TB for TB detection

The mean Area under the Receiver Operating Curve (AUC-ROC) of the ensemble model (AID.TB) for predicting TB on the unseen test set is 0.79 (95% CI: 0.73–0.85), as seen in Figure 3. At a sensitivity of 89.9% (95% CI: 82.4%–94.4%), the ensemble model has a specificity of 50.4% (95% CI: 42.0%–58.7%) for TB.

**Figure 3:**
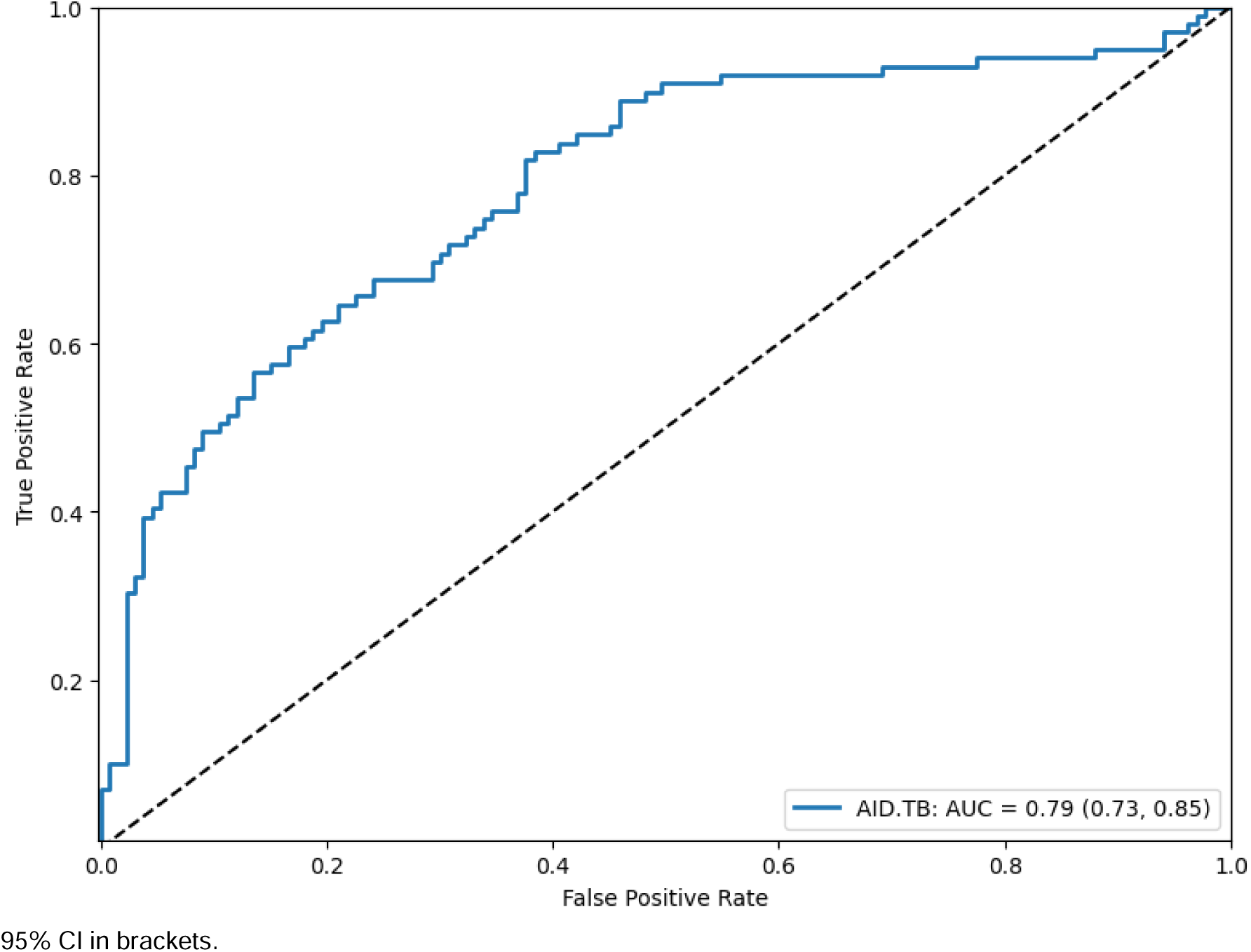
AUC-ROC for AID.TB performance on the hold-out test set.

For the threshold derived from the fold validation set at ∼90% sensitivity the specificity is 56.7% (± 4.9%). Using this same threshold on the hold-out test set yielded a sensitivity of 81.0% (± 4.2%) and a specificity of 53.9% (± 2.5%) for TB.

### Performance of AID.TB for TB detection in key subgroups

**Table 2** summarizes the average performance of AID.TB ensemble model on the test set (n=233) in key subgroups. The model had similar AUCs on the test set in participants that are HIV positive and HIV negative, with AUCs of 0.79 (95% CI: 0.68–0.90) and 0.79 (95% CI: 0.72– 0.86) respectively. In HIV positive participants (n=72), the model has a sensitivity of 86.1% (95% CI: 71.3%–93.9%) with a corresponding specificity of 22.2% (95% CI: 11.7%–38.1%), and for HIV negative participants (n=161) a sensitivity of 87.3%% (95% CI: 76.8%–93.4%) with corresponding specificity of 52.6% (95% CI: 42.7%–62.2%).

**Table 2:** AID.TB performance on the hold-out test set in key subgroups.

| Subgroup | n | AUC (95% CI) | Sen % (95% CI ) | Spe % (95% CI ) |
| --- | --- | --- | --- | --- |
| HIV Pos | 72 | 0.79 (0.68-0.90) | 86.1% (71.3%–93.9%) | 22.2% (11.7%–38.1%) |
| HIV Neg | 161 | 0.79 (0.72-0.86) | 87.3% (76.8%–93.4%) | 52.6% (42.7%–62.2%) |
| Symptomatic | 194 | 0.79 (0.72-0.85) | 89.4% (82.5%–94.1%) | 49.0% (39.4%–58.6%) |
| Asymptomatic | 38 | 0.67 (0.35-0.93) | 80.0% (37.6%–96.4%) | 27.3% (15.1%–44.2%) |

For participants with a positive TB symptom-screen (n=194), the AUC was 0.79 (95% CI, 0.72– 0.85), with a sensitivity of 89.4% (95% CI: 82.5%–94.1%) and corresponding specificity of 49.0% (95% CI: 39.4%–58.6%). For participants that were asymptomatic, i.e. did not have TB symptoms (n=38), the model achieved an AUC of 0.67 (95% CI, 0.35–0.93), with a sensitivity of 80.0% (95% CI: 37.6%–96.4%) and corresponding specificity of 27.3% (95% CI: 15.1%–44.2%).

## Discussion

At a sensitivity of 89.9% (95% CI: 82.4%–94.4%), this AI model (AID.TB) trained on digital lung auscultation sounds achieved a specificity 50.4% (95% CI: 42.0%–58.7%) for pulmonary TB in this primary care patient population in the Western Cape, South Africa.

According to the 2024 WHO target product profiles (TPPs) for TB diagnostics, a triage or referral test should achieve a minimum sensitivity of 90% and specificity of 70%. While AID.TB meets the sensitivity threshold, its specificity falls below this standard.^3^ AID.TB’s performance in this study does not yet meet the current WHO requirements for a triage or referral test.

Several factors relating to the clinical presentation of TB and other pulmonary diseases may impact the model’s performance. First, overlap exists in the acoustic patterns of many pulmonary diseases. As an example, inspiratory crackles are common clinical findings in both TB pneumonia, non-TB pneumonia, and bronchiectasis. Similar overlap also occurs in participants with prior TB and persistent lung damage. In addition, abnormal lung sounds in TB positive participants vary depending on the stage of presentation (early vs. late) and in HIV coinfection. Lastly, the presence of significant background noise may mimic abnormal lung sounds and reduce model performance.^8^

In this study, all participants were suspected to have TB, and 83% (n=970) participants had at least one TB symptom. Many participants that tested negative for TB on Xpert Ultra may have had respiratory illness other than TB, but with similar abnormal lung sounds to TB. As a result, differentiating cases and controls in this study is a diagnostic challenge, and hence contributed to the model’s performance.

Importantly, enrolling participants suspected to have TB reflects the real-world application of TB screening tools. Had the study recruited only healthy participants as controls and confirmed TB cases, the AID.TB model performance and specificity would be higher but at the cost of generalizability to real-world screening settings.

### AID.TB performance compared to the current WHO screening guideline

AID.TB has comparable performance to all other TB screening methods in the WHO’s 2024 screening guideline.^2^ Notably given the sensitivity and specificity results above, the AID.TB ensemble model outperforms 3 out of 12 of the CAD-CXR technologies included in a 2024 independent review of CAD-CXR for TB in a similar study population in South Africa (Figure 4).^14^

**Figure 4:**
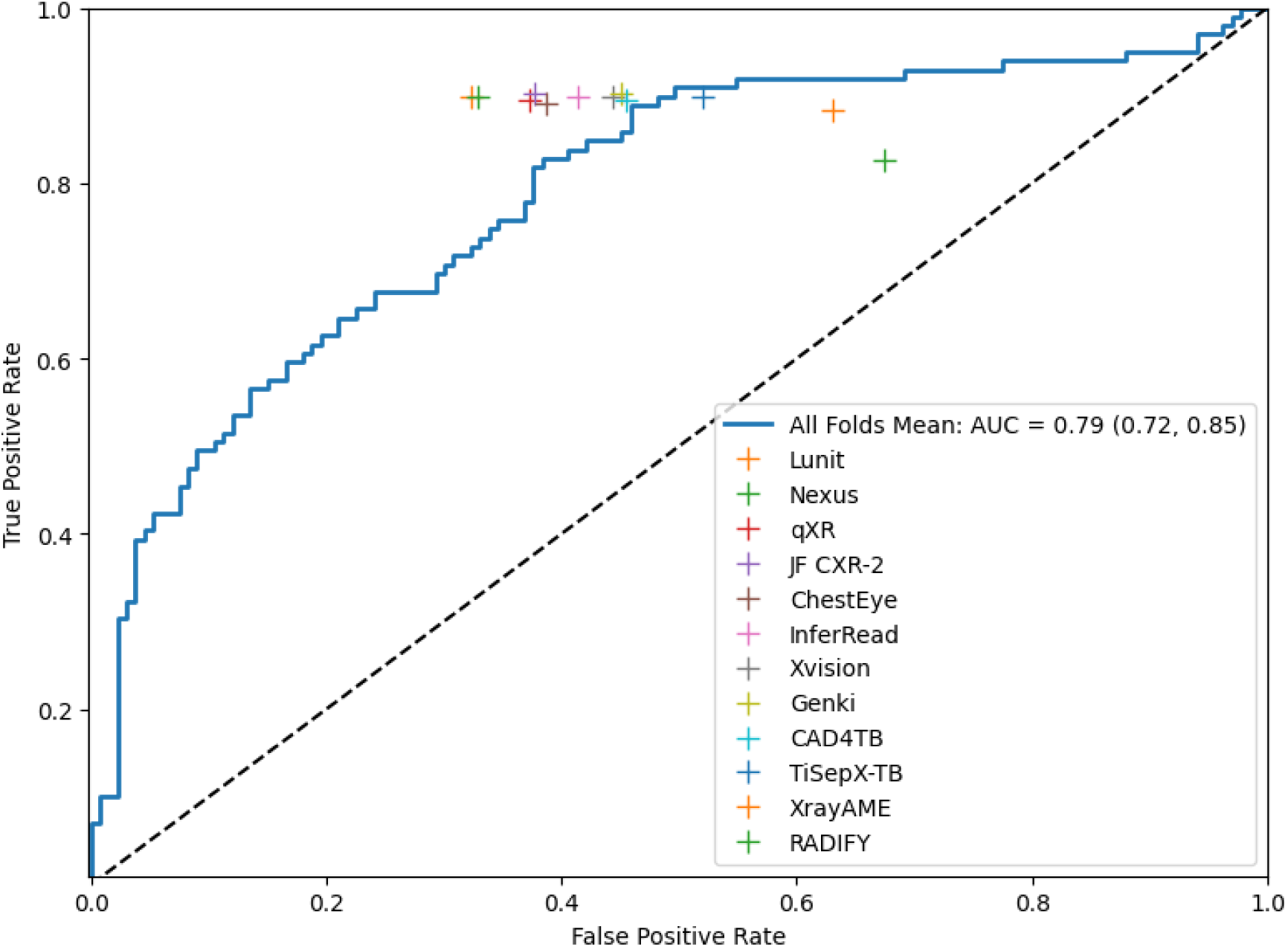
AID.TB AUC-ROC compared to 12 CAD-CXR technologies evaluated in South Africa. *Note: the CAD-CXR performance results are included in the above figure for comparison only. These technologies were evaluated as part of a large external validation study performed in a similar South African population, published September 2024.^12^

It is important to note that although the population is somewhat similar to the one in this study, these are two different studies performed at different times and hence this is an indirect comparison. The main difference between the two study populations is the HIV coinfection rate, which is 31% in this study and 18% in the CAD-CXR external validation study.^14^

AID.TB also has comparable performance to clinicians interpreting CXR for TB. However, the performance of clinician interpretation of CXR for TB is highly variable. This is largely impacted by the level of experience of the clinician. According to the 2016 WHO guideline on CXR for TB detection, CXR interpretation by clinicians for TB has a sensitivity ranging between 87% and 98% and specificity between 46% and 67%.^15^

AID.TB significantly outperforms symptom-based screening, which has the worst performance of the available screening methods, with a sensitivity of 71% and specificity of 64% according to the WHO’s 2021 guideline for TB screening.^2^ Importantly, a 2020 review of TB prevalence surveys in 24 countries found that 50.4% (IQR=39.8%–62.3%) of TB positive cases were asymptomatic.^4^ Symptom-based screening is still widely used globally due to the lack of access to more effective TB screening methods, and so a significant proportion of global TB cases are missed.

### AID.TB performance in subgroups

It is important to note that the sensitivity and specificity results for subgroups were limited by the small sample sizes of subgroups in the hold-out test set.

A similar model AUC was observed in HIV positive and negative participants (AUC=0.79) but with a larger CI in HIV positive participants (95% CI: 0.68–0.90), **Table 2**. Of note is the significantly lower specificity in HIV positive versus negative participants (22.2% vs. 52.6%), at thresholds with similar sensitivity. This likely reflects the atypical presentation of TB in HIV coinfection, particularly in advanced disease (CD4 < 200 cells/mm^3^).^16^

A compromised immune system responds less to the TB bacillus. Due to this, these patients typically have less pulmonary consolidation and cavitation compared to HIV negative patients.^16^ As a result, participants with advanced HIV present with fewer abnormal findings on auscultation which impacts model performance.

In contrast, HIV positive participants that are well controlled (CD4 > 500 cells/mm^3^) are likely to present similarly to immunocompetent individuals.^16^ Studies on CAD-CXR for TB have produced similar findings, showing a lower AUC in HIV positive participants compared to HIV negative participants.^14^ For these reasons, future research should include CD4 count as this would be a more accurate guide for atypical presentation.^17^

AID.TB showed some utility in the asymptomatic group (AUC=0.67, 95% CI: 0.42–0.93), however the number of TB positive participants in this group was small (n=38). This limits the interpretability of this result and will require further research.

### Strengths and limitations

This study has several strengths. The first is the high HIV prevalence within the training dataset (31%), resulting in improved model generalizability to other populations with a high HIV prevalence. Similarly, the high TB prevalence in the patient population contributes to improved model generalizability in other populations with a high TB prevalence.

Despite the case-control study design and risk of selection bias, the participants in this study represent a heterogenous group due to the way participants were recruited (after reporting having had a sputum Xpert test). This inclusion of presumptive TB cases included both symptomatic and asymptomatic at-risk patients which also improves generalizability.

Another strength of the study is the single reference standard used, namely sputum Xpert Ultra.

Lastly, the study showed that a digital stethoscope and AI model can be used effectively for TB screening by nurses in a primary healthcare setting.

The main limitation of the study is that the study population is South African only and therefore model generalizability to other populations needs to be validated.

The case-control study design is a limitation, as it may result in optimistic estimates of performance due to potential selection of extreme cases, i.e. spectrum bias. This may inflate model performance. However, the risk of spectrum bias in this study is likely low. All participants were recruited based on clinical suspicion of TB, and 83% had at least one TB symptom. As a result, the study population had similar clinical presentations rather than at the extremes of the disease spectrum.

The risk of model bias across demographic subgroups was not formally assessed and is a limitation. Future research should include subgroup analyses with larger subgroup sample sizes to assess fairness.

Importantly, digital chest auscultation at 6 chest positions covers less of the chest area than the 10-12 positions usually performed during clinical examination and is a limitation of this study. This loss in lung auscultation data may result in abnormal auscultation findings being missed which would reduce model performance during screening.

Lastly, the study did not directly compare the performance of human auscultated lung sounds (normal vs abnormal) or directly compare digital stethoscope performance against CXR with a human interpreter or CAD technology.

## Conclusion

This AI model, AID.TB, trained on lung sounds from auscultation performed by nurses using a digital stethoscope, shows early promise as a method for TB screening. In addition, the digital stethoscope is portable, with low hardware costs, and is easy to use after basic training. If validated, it would align well with the WHO’s End TB Strategy.

Compared to symptom-based screening, this method could significantly reduce missed TB-positive cases. It shows promising performance when compared to CAD-CXR after training on significantly less data than CAD-CXR models.^18^ The performance of the AID.TB model is expected to improve after training on larger data volumes and diversity. Similar model performance improvements were observed in CAD-CXR after training on larger data volumes.^19^

Future research should expand the database of abnormal TB lung sounds to improve model performance and should focus on prospective diagnostic accuracy studies and validation in diverse populations. There should be emphasis on both high and low HIV-prevalence settings as well as on asymptomatic TB cases and active case-finding.

Further research should also be undertaken by groups independent of the product developer, and include data on costs, cost-effectiveness, and usability. A direct head-to-head comparison of accuracy and cost-effectiveness of TB screening with the AI-based digital stethoscope versus screening with CAD-CXR would also be valuable.

## Data and code availability

### Data availability

The data used in this study are proprietary and were collected as part of research conducted by AI Diagnostics. As such, they are not publicly available. Requests for data access will be considered on a case-by-case basis and will require a data use agreement and approval.

### Code availability

The code used for model evaluation and statistical analysis may be shared upon reasonable request, subject to review and approval by AI Diagnostics.

### Ethics approval and informed consent

Ethics approval for this study was obtained from the Pharma- Ethics, South Africa. Protocol number: AID-CIP-001, reference number 201023634, 27 November 2020. The study protocol can be made available upon reasonable request.

Written informed consent was obtained from all participants prior to enrolment. For participants under the age of 18, consent was obtained from a parent or legal guardian, and assent was obtained from the participant where appropriate. The study was registered with the Provincial Health Research Committee (PHRC) of the Western Cape, South Africa, refernece number: WC 202102 010.

### Contributions

From authors:

Dr Max Rath: led manuscript development, interpretation of results, discussion, strength and limitations, and conclusion. Provided input into data preparation, statistical analysis and model training and evaluation.

Johan Coetzee: assisted with protocol development, led technical support, data preparation, statistical analysis, and model training and evaluation.

Mark van Breda: assisted with protocol development, provided operational and technical support during data collection.

Braden van Breda: led as principal investigator. Led protocol development and data collection. Provided oversight and input into data preparation, statistical analysis, model training and evaluation, results interpretation, and discussion.

From non-authors:

Luca Powell (AI Diagnostics): assisted with statistical analysis.

Prof Madhukar Pai (McGill University): manuscript draft review and feedback.

Dr Jeremy Nel (University of the Witwatersrand): manuscript draft review and feedback.

AI tools, namely ChatGPT and Microsoft Copilot were used for research and literature search, as well as for grammar, spelling, and formatting review.

